# Development of a Research Program Focused on Non-Invasive Brain Stimulation for Peripheral Neuropathy in Minoritized Communities

**DOI:** 10.1101/2025.01.10.25320213

**Authors:** Marlon L. Wong, Lisa M. McTeague, Chelsea A. Miller, Gabriel Gonzalez, Melissa M. Tovin, Frank J. Penedo, Eva Widerstrom-Noga

## Abstract

Black and Hispanic/Latino communities experience disproportionate chronic pain and are underrepresented in pain research. Transcutaneous auricular vagus nerve stimulation (taVNS) and transcranial magnetic stimulation (TMS) are promising tools for pain management. Therefore, it is critical to ensure that research using these tools engages underrepresented communities to make research findings more generalizable and reach all who may benefit. Lack of diversity in the research workforce itself is a key barrier to improving Black and Hispanic/Latino representation in pain research, and video-enhanced recruitment and consenting may be a useful tool to better engage minoritized communities.

Using community participatory research principles in an iterative process, we engaged key stakeholders, including neuromodulation researchers and minoritized community members, to create and test informational videos on taVNS and TMS. These videos were designed for Black English-speaking, Hispanic/Latino Spanish-speaking, and Haitian-Creole speaking people with chronic pain. Study 1 involved iterative feedback from stakeholders to develop test videos, which were then refined based on community member input. Study 2 was a pilot randomized controlled trial assessing the impact of these videos on participant expectations for pain relief with taVNS.

Results indicated that the videos were well-received, and there was no significant difference in expectancy scores between those who viewed the videos and those who received traditional brochures. This suggests that while videos may improve engagement, they do not unduly influence expectations, potentially making them valuable tools for improving research participation in underrepresented populations. These videos will be freely available to help researchers to engage people from minority communities.

**PERSPECTIVE:** This article presents the process of developing culturally sensitive informational videos on taVNS and TMS, and provides the field with these videos in English, Spanish, and Haitian-Creole language. These videos could potentially help researchers to engage people from minority communities to enhance the diversity and reach of research using noninvasive brain stimulation for pain.

## INTRODUCTION

Black and Hispanic/Latino communities are disproportionately affected by chronic pain, with greater prevalence and worse outcomes than non-Hispanic White communities, yet these same communities are underrepresented in pain research studies.^1-5^ Transcutaneous auricular vagus nerve stimulation (taVNS) and transcranial magnetic stimulation (TMS) are forms of non-invasive brain stimulation (NIBS) with great promise for improving pain management. In fact, there is rapidly growing interest in taVNS and TMS, as demonstrated by the >300 clinical trials currently investigating these interventions for pain (listed on clinicaltrials.gov). However, under-representation of Black and Hispanic/Latino communities is particularly problematic in NIBS research due to a number of factors, including lack of diversity in the research workforce, incompatibility between the technologies and certain hair types and styles, and reluctance of minoritized populations to participate in clinical trials.^6^ It is critical that research on taVNS and TMS includes perspectives of people from minoritized communities to ensure that the research reaches all of those who may benefit and to make research findings more generalizable. Achieving greater diversity in the NIBS research workforce is an important long-term goal for improving this problem. In the present, however, meaningful short-term efforts can be undertaken to improve the inclusion of participants representative of the overall population within NIBS research studies.

A commonly cited barrier to recruiting minoritized people for research is medical mistrust.^7^ The best known approach for overcoming this barrier is community-engaged research,^8^ and large centers with a history of community-engaged research have been found to offer better opportunities for racially minoritized groups to participate in research.^8^ Unfortunately, community-engaged research methods can be challenging to employ, particularly for smaller institutions with limited resources.

Video-enhanced recruitment and consenting (VE-RC) can be used to better engage minoritized communities, bridging the potential racial discordance between researchers and minoritized participants. Although the results of a Cochrane review in 2014 suggested that VE-RC made little difference in the rate or willingness to participate, the authors acknowledged that the value of VE-RC was unclear, and they encouraged investigators to continue exploring VE-RC.^9^ In fact, studies published since then consistently indicate that VE-RC improves participant satisfaction^10, 11^ and improves understanding and retention of the information provided.^12-14^ This is not surprising, as it is widely recognized that many learners experience more effective communication in visual formats such as video. In one of the largest studies on the topic, Fanaroff et al.^15^ compared the enrollment performance of text only sites to sites with VE-RC in a multicentered study that included 7,904 patients at cardiology, endocrinology, and primary care clinics across the United States. Compared with text only sites, VE-RC sites enrolled more patients ≥75 years old, more Black patients, and more patients without college degrees. Indeed, a recent systematic review on VE-RC in cancer trials concluded that “video interventions are well-received by [minoritized] survivors and may improve [minoritized] representation in clinical trials, yet they are underutilized.”*^10^*

Information provided in video format is often perceived as more credible,^16, 17^ and when done from an inclusive perspective, may help to engender better recruitment and retention rates.^10, 15^ Such videos may be used for informing communities, recruiting participants, and enhancing the consenting process. However, the development of informational videos must be done with care and with the participation of the communities themselves to avoid missteps that could cause further alienation. Thus, this study has three primary goals: 1) involve Black English-speaking, Hispanic/Latino Spanish-speaking, and Haitian Creole-speaking communities, to develop informational videos on taVNS and TMS; 2) make these videos publicly available to provide informational materials for the field at large; and 3) establish infrastructure and community relationships for a research agenda focused on equitable and inclusive research on NIBS for pain management.

## METHODS

We used a “technical-scientific and positivist”^18^ model of action research, with the intention of linking research to action. In this approach the research questions and theoretical resources were set independent of interaction with community members, and participants acted as “on the ground feedback.”^18^ By researching and developing culturally sensitive videos, our intention was to provide ourselves, and the field, with data and tools to enhance racial and ethnic diversity in NIBS research. This project consisted of 2 studies. First, we used an iterative process to engage key stakeholders for input and feedback in the video development process (Study 1, IRB# 20230210). Then, we tested the videos in a pilot randomized controlled trial (Study 2, clinicaltrials.gov #NCT05896202). For both studies, approval was granted by the University of Miami Institutional Review Board, and informed consent was obtained from participants.

### Study 1

A generic qualitative research design, from an interpretivist paradigm, was employed for this project. This design was chosen because it would allow for the researchers to adapt their methods to fit the specific needs of their study as the project unfolded.^19^ Key stakeholders, including neuromodulation researchers, recruiters, and racial/ethnic minoritized community members with chronic pain, were engaged in an iterative process throughout video development (**Figure 1**). To get input from neuromodulation practitioners, researchers from the National Center of Neuromodulation for Rehabilitation (NC-NM4R) were first engaged via one-on-one interviews and two focus groups. Input from a small focus group of researchers at the Berenson-Allen Center for Noninvasive Brain Stimulation was also obtained during the second round of the feedback process. For minoritized community member input, individuals with chronic pain from the representative communities were recruited locally in the Miami-Dade County area through health fairs, flyers in clinics, and snowball sampling to purposefully identify potential participants. Participation was in-person or remote via Zoom, determined by preference.

**Figure 1.**
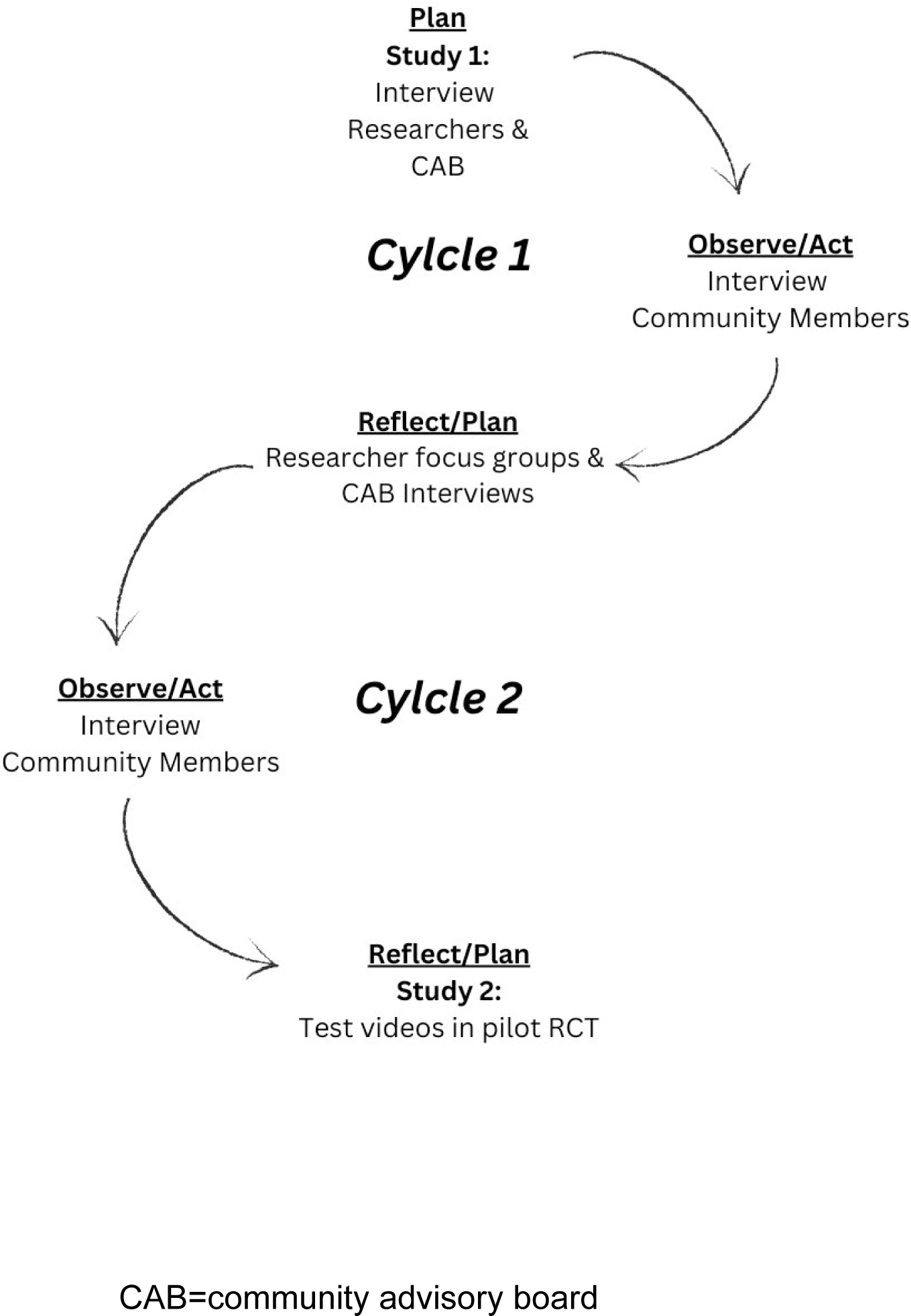
Iterative data Collection Process Embedded within the Action Research Cycle.

### Neuromodulation researchers

As the first step in the project, 60-minute informal and semi-structured one-on-one interviews were conducted with 10 researchers from the NC-NM4R, and a large 90-minute focus group of NC-NM4R researchers and recruiters was conducted by the principal investigator (MW), who did not have prior relationships with the interviewees. The interviews and focus group were used to generate ideas for the video content and the desired mood or feel of the video to make sure that the views of those conducting NIBS research were represented in the end-product. The format was deliberately informal (i.e., no audio recordings) and semi-structured, to better facilitate idea generation, open feedback, and to encourage diverse perspectives. A sample mood reel was presented to stimulate ideas and feedback, which consisted of a 60-second video with sample footage of investigator-participant interactions, animations of neurophysiological effects, and on-screen text. The following questions were used to guide the discussion:

- What are your challenges with recruiting and retaining Black and Hispanic/Latino participants for your research?
- How could videos best be used to help improve the recruitment and retention of Black and Hispanic/Latino participants?
- What content do you think should be in a video?
- Describe how you envision a short animation would be used to best describe the underlying mechanisms.
- Here is a sample framework for the videos. What are your thoughts?

Using the feedback from researchers, test videos were then developed, and these test videos were presented to NC-NM4R researchers via two additional informal and unstructured small focus groups for feedback. Feedback was also received from a small focus group of researchers at the Berenson-Allen Center for Noninvasive Brain Stimulation. Field notes were made during and after the interviews and focus groups.

### Community Member Participants

Two representatives from each ethnic/racial group of interest were recruited for the community advisory board (CAB), for a total of 6 CAB members. CAB members reported strongly identifying with their respective communities, reported that English, Spanish, or Haitian-Creole were their primary language, and agreed to meet twice from 6/2023 to 5/2024 to provide feedback on the research plan, recruitment strategy, test videos, and the interim summary findings and interpretations. Four of the CAB members also had chronic pain.

All other community member participants were recruited for a single session interview, and they were required to meet the following criteria: 1) be at least 18 years old, 2) have persistent pain of any etiology (e.g., back pain, diabetic neuropathy, knee arthritis) for >3 months, and 3) self-identify as one of the target communities and reported that their primary language was English for the those in the Black English-speaking group, Spanish for those in the Hispanic/Latino group, or Haitian-Creole for those in the Haitian-Creole speaking group. Interviews and focus groups were conducted with racial/ethnic concordant investigators in the preferred language for each participant.

Specifically, all Black English-speaking participants were paired with a Black English-speaking investigator, Spanish-speaking participants were paired with a native Spanish-speaking investigator, and Haitian-Creole speaking participants were paired with a native Haitian-Creole speaking investigator. The interviews and focus groups were audio recorded and transcribed, and non-English recordings were translated into English by a certified professional translation service (GMR Transcription) for analyses. The first round of semi-structured interviews was used to gain feedback on the taVNS test videos. Then focus groups of only the CAB members were conducted to provide member checking on interpretation of the interview findings.

In addition to interview or focus group participation, all community member participants completed the following questionnaires (in their preferred language):

*Adapted Group-Based Medical Mistrust Scale (MMS)*^20^: Medical mistrust has been widely reported as a key barrier to participation of minoritized persons in clinical trials.^21^ The Group-Based Medical Mistrust Scale was originally developed, and has been widely used, to assess medical mistrust in minoritized people who contact the health care system.*^22^*The original Group-Based Medical Mistrust Scale was adapted to measure mistrust in health researchers (MMS), and the adapted version has also been shown to have strong psychometrics.^20^ Thus, the MMS was used in this study to assess community member participants’ degree of mistrust in medical research. The MMS is a 6-item questionnaire assessing a person’s beliefs that their race/ethnic group is prone to mistreatment in medical research. Each item is scored on a 5-point Likert scale, ranging from strongly disagree (1) to strongly agree (5). We report MMS scores as a total of the 6 items, with the last item reverse scored and potential scores ranging from 6-30, and higher scores indicate greater mistrust in medical research.

*Credibility/Expectancy questionnaire Version II (CEQ) ^23^*: Participants’ feelings of uncertainty pertaining to the credibility and expectancy of researchers and the interventions being studied has also been reported as a barrier to minoritized participation in clinical trials. Thus, credibility and expectancy for pain relief was assessed using the CEQ. The CEQ is a validated tool for assessing participants’ perceptions on the credibility of therapeutic tools and their expectations for symptom improvement. The questionnaire consists of 6 questions, scored on a Likert scale ranging from 1= Not at all useful to 9=Very useful (with 5 representing “Somewhat useful”). We report the mean of the 6 items as the CEQ scores, with higher scores indicating greater perceived credibility of taVNS and expectancy for pain relief with taVNS.

*Protocol for Responding to and Assessing Patients’ Assets, Risks, and Experiences (PRAPARE)^24^*: Social determinants of health are known to influence health care utilization and outcomes. The PRAPARE is a nationally standardized, and widely used, screening tool for assessing an individual’s social drivers of health. It consists of 21 items covering the domains of personal characteristics, family and home, money and resources, social and emotional health, and institutional or environmental vulnerability (i.e., recent time spent in jail/prison, refugee status, and spousal abuse). In this study, the PRAPARE was used to gather important contextual information on the participants in this study, but it was not scored.

The MMS and PRAPARE were administered at the beginning of each session, and the CEQ was administered after participants viewed the videos and provided feedback.

### Study 2

The pilot was a single-blinded, sham-controlled, feasibility study that was designed to examine the influence of the newly developed videos on participant expectations for pain relief with taVNS in Black and Hispanic/Latino people with chemotherapy induced peripheral neuropathy (CIPN) or diabetic neuropathy (DN). A target sample size of 24 was chosen for this study because it was estimated that this would provide the needed power to reach saturation with qualitative analysis.

The pilot RCT included only Black and Hispanic/Latino patients with CIPN or DN, and block randomization by race/ethnicity was used to ensure that there was equal representation of these groups across the intervention and control groups. Due to financial constraints, we were unable to accommodate Haitian-Creole speakers in study 2. Participants were recruited from the University of Miami medical health care system from January to May 2024. Potential participants were identified by medical record and then their respective providers (i.e., oncologist or endocrinologist) informed them about the pilot study during clinical visits. Inclusion criteria included anyone with glove or stocking distribution paresthesia or dysesthesia that developed after receiving neurotoxic chemotherapies or with a diagnosis of diabetic neuropathy and who self-identified as Black or Hispanic/Latino. Exclusion criteria included 1) any unstable medical condition or medical contraindication to moderate physical exertion (e.g., unstable angina or cardiac arrythmia), 2) pregnancy, 3) presence of cognitive impairment or language barrier that impairs full autonomy in the consent process or in the ability to participate in detailed interviews, 4) implants in the head or neck, cochlear implants, or pacemaker, 5) head or neck metastasis or recent ear trauma, 6) history of seizures.

Participants were randomly assigned to video or control groups, and all participants completed 3 visits. The first visit consisted of ∼90 minutes of education on taVNS, including review of brochures and consent forms (both groups) and 3 short video segments on taVNS for the intervention group. The videos contained the same content as the brochures and consent forms, so all participants received the same information but in different formats. Further, all participants had ample opportunity to ask questions and discuss the content with the investigators. Racial and ethnic differences between participant and investigators/providers is also known to influence expectations and pain outcomes;^25, 26^ thus, a Black investigator provided all educational sessions for Black participants, and a Hispanic/Latino investigator provided all education sessions for Hispanic/Latino participants and in their preferred language (English or Spanish). Both investigators provided the same information to participants. At the end of the educational session, participants provided feedback on the educational materials and completed the EXPECT^27^ questionnaire. The EXPECT is a 4-item questionnaire that assesses expectations for pain improvement. Each of the 4 items is scored on an 11-point scale, with 0 being no change and 10 representing complete relief.^27^ We hypothesized that participants who viewed the videos would have higher EXPECT scores.

### Quantitative analysis

For both study 1 and study 2, descriptive statistics were assessed for demographic data and questionnaire findings. This included sample means, medians, and standard deviations for each continuous variable, and frequencies and percentages for categorical variables. Group comparisons were conducted using Krustal-Wallis and Chi-Square tests, and correlations were assessed using Spearman’s rho. All statistical analyses were conducted using Statistical Package for the Social Sciences (SPSS) v29 (IBM Corp., Armonk, NY) and figures were rendered using GraphPad Prism v10.4.0 (Graphpad Software, La Jolla, CA).

### Qualitative analysis

Rapid qualitative analysis is widely used for implementation projects, such as this, when the goal is to create change in response to the findings rather than to generate new theories. Rapid qualitative analysis was systematically applied in this study according to established protocols.^28^ The interview guide was used to create structured templates and matrix displays to facilitate data condensation, synthesis, and theme development. Templates were developed collaboratively by the team (MW, CM, CG) and pilot tested to ensure consistency, usability, and relevance. Once consistency was demonstrated, MW completed summaries of transcripts from sessions with Black English-speaking participants, both GG and CM completed summaries of transcripts for Spanish-speaking participants, and CM completed summaries of transcripts for Haitian Creole-speaking participants. The summaries were aggregated by MW and CM to populate the matrices that enabled systematic comparison between participants.

All three team members who were engaged in the qualitative analysis were physical therapists and pain-scientists, with an interest in health equity, and with extensive experience working with the target populations in clinical settings. As mentioned earlier, all recordings were transcribed in English, and the racial and ethnic backgrounds of the team members who analyzed the transcripts were Black Caribbean (MW, male), White American (CM, female), and Latino Mexican (GG, male). MW had experience with qualitative and mixed-methods research through involvement in several funded projects, and he received formal mentorship in qualitative research from recognized experts as part of the pilot funding programs that supported this project. CM had completed formal coursework and training in qualitative research as part of attaining her master’s degree in public health. This was GG’s first exposure to qualitative research, and he received training in interviewing and qualitative research principles prior to initiating data collection processes.

## RESULTS

### A. DEVELOPMENT of TEST VIDEOS

#### A.1 Neuromodulation Researchers’ Initial Input for Video Content

A variety of perspectives were gained from the informal interviews and focus group with NC-NM4R researchers. Most felt that the best use of the videos would be for recruitment purposes, while some felt that they would best help with enhancing the consent process, and a few felt that the best use of videos would be to direct them to the researchers, enhancing their cultural sensitivity. Recruitment was commonly viewed as the biggest barrier. Thus, we decided to design the videos to be primarily informational, to assist with recruitment efforts.

Opinions also differed on the optimal length and content for the videos, with some advocating for very brief videos (less than 1 minute) to capture attention and generate interest with recruits, and others advocating for more lengthy videos (over 5 minutes) to provide detailed information on the research procedures as well as the risks and regulations involved. As a result, we decided to start with the taVNS content and create test versions for full length videos (∼6 minutes in length) as well as segmented versions, in which the full-length videos were divided into 3 separate video segments organized around the following content areas: 1) introduction and how taVNS works, 2) risks and regulations, and 3) what to expect when you participate in taVNS research (**Table 1**). In the introduction segment, information on the potential indications for taVNS were provided, and an animation of the proposed mechanisms of taVNS was presented. In the risks and regulations segment, the potential side effects were outlined, and the process of research oversight was described. In the final segment, entitled *what to expect*, the process of prepping and applying taVNS was demonstrated. We used block allocation, with alternating assignment within each racial/ethnic group (i.e., the first participant received the full-length version, and the second participant received the segmented version for the Black, English-speaking group), to get feedback on the full length and segmented versions of the videos.

**Table 1.**
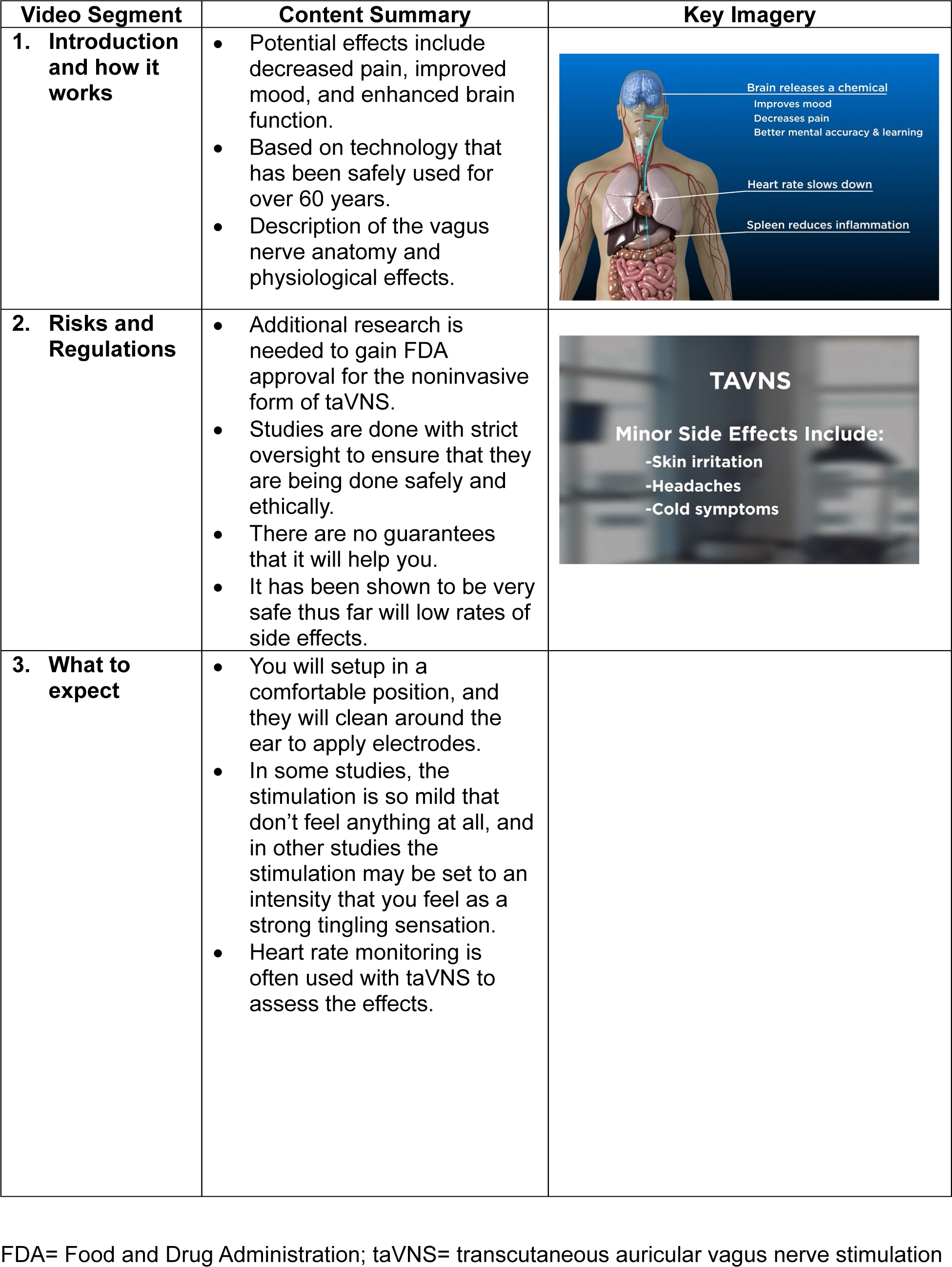
Overview of Test Video Segments.

| Video Segment | Content Summary | Key Imagery |
| --- | --- | --- |
| 1. Introduction and how it works | Potential effects include decreased pain, improved mood, and enhanced brain function.<br>Based on technology that has been safely used for over 60 years.<br>Description of the vagus nerve anatomy and physiological effects. |  |
| 2. Risks and Regulations | Additional research is needed to gain FDA approval for the noninvasive form of taVNS.<br>Studies are done with strict oversight to ensure that they are being done safely and ethically.<br>There are no guarantees that it will help you.<br>It has been shown to be very safe thus far with low rates of side effects. |  |
| 3. What to expect | You will setup in a comfortable position, and they will clean around the ear to apply electrodes.<br>In some studies, the stimulation is so mild that don't feel anything at all, and in other studies the stimulation may be set to an intensity that you feel as a strong tingling sensation.<br>Heart rate monitoring is often used with taVNS to assess the effects. |  |

FDA= Food and Drug Administration; taVNS= transcutaneous auricular vagus nerve stimulation

#### A.2 Community Member Participants: Round 1

Sixteen community members, including both the CAB and community participants with chronic pain, provided feedback on the test videos produced in their respective languages (5 Black English speaking, 6 Spanish Speaking, and 5 Haitian-Creole speaking). The group mean (standard deviation [SD]) for age was 51.6 (16.2) years, and 12 (75%) of the participants were female. The 14 participants with chronic pain had moderate pain on average with mean (SD) scores, on a scale of 0-10, for current, best, and worst pain intensities within the last week of 4.9 (3.2), 3.0 (2.7), and 6.9 (3.4), respectively. There were no significant differences between racial/ethnic groups in other demographics, pain characteristics, or social risk factors (**Table 2**). The group was diverse in social risk factors, with 3 of the participants not having any health insurance, 3 with Medicaid coverage, 5 with Medicare, and 5 with private insurance. Four of the participants reported that within the last year, they or a family member they live with were unable to get medicine when it was needed. There was also diversity in medical mistrust, with a group mean (standard deviation) score on the MMS of 17.8 (6.6) (median =18.5) and scores ranging from 6-26.

**Table 2.** Pain Characteristics of the Community Participants.

| Primary Language | Sex | Diagnosis | Current Pain | Best Pain | Worst Pain | Pain Status |
| --- | --- | --- | --- | --- | --- | --- |
| Round 1 |  |  |  |  |  |  |
| English | Female | Knee pain | 6 | 2 | 8 | Not changing |
| English | Female | Lumbar degenerative joint disease | 3 | 3 | 10 | Getting worse |
| English | Male | Postherpetic neuralgia | 2 | 1 | 6 | Getting better |
| English | Female | Spinal Stenosis | 8 | 9 | 2 | Getting worse |
| English | Female | Arthritis | 0 | 0 | 7 | Coming and going |
| English | Female | Arthritis | 7 | 3 | 10 | Not changing |
| Spanish | Male | --- | --- | --- | --- | --- |
| Spanish | Female | Lumbar herniated disc | 10 | 8 | 10 | Getting worse |
| Spanish | Female | Migraine | 9 | 2 | 8 | Getting worse |
| Spanish | Female | Scoliosis | 6 | 3 | 10 | Coming and going |
| Spanish | Male | Back pain | 7 | 3 | 10 | Coming and going |
| Haitian-Creole | Female | --- | --- | --- | --- | --- |
| Haitian-Creole | Female | Lumbar radiculopathy | 4 | 2 | 7 | Coming and going |
| Haitian-Creole | Female | Abdominal pain | 4 | 1 | 8 | Not changing |
| Haitian-Creole | Male | Angina | 6 | 5 | 8 | Coming and going |
| Haitian-Creole | Female | Arthritis | 6 | 4 | 6 | Coming and going |
| Round 2 |  |  |  |  |  |  |
| Spanish | Female | Lateral epicondylitis | 4 | 3 | 9 | Coming and going |
| English | Female | Osteoarthritis,<br>Congenital spinal<br>stenosis | 10 | 1 | 10 | Getting worse |
| Spanish | Female | Low back pain | 5 | 3 | 8 | Not changing |
| Haitian-Creole | Female | Foot pain | 4 | --- | --- | Not changing |
| English | Female | Plantar fasciitis | 5 | 3 | 10 | Getting worse |
| Haitian-Creole | Male | Headaches | 0 | 0 | 0 | Coming and<br>going |
| Haitian-Creole | Female | Knee pain | 5 | 3 | 8 | Getting worse |
| English | Female | Ankylosing spondylitis | 7 | 5 | 10 | Coming and<br>going |
| Spanish | Female | Shoulder pain | 6 | 5 | 8 | Not changing |

The interviews identified several potentially important differences across racial/ethnic groups for participants’ current pain management strategy. All Black, English-speaking participants used medication as their primary pain management strategy, and none had ever tried electrical stimulation. Conversely, 4 of the Spanish-speaking participants (80%) used medication for pain management when needed, two had tried electrical stimulation (40%), and all reported cognitive/emotional or active coping strategies (i.e., distraction, “learning to live with it,” breathing, going to the gym) as primary forms of pain management. Three of the Haitian Creole-speakers (60%) reported using medication for pain when needed and all reported using alternative or “natural” treatments (i.e. tea, massage, oil) as their primary pain management approach.

#### A.3 Community Member Participant Feedback on Test Videos

Participants watched the videos in their primary language and provided feedback in their primary language (i.e., American English, Caribbean Spanish [since these videos were intended to be used in Miami-Dade County], and Haitian-Creole). Overall, the videos were well received by participants, and details on their feedback and the decision log can be found in **Supplement 1**. Key themes identified across all groups were that they most appreciated the animation and education on “how it works”, they found the video to be “clear”, and there was concern about taVNS not being FDA approved. The general appreciation for the videos’ educational value is best articulated by the following participant comment:

*“This is a positive all the way across the board, because I can tell you it by being a black female and dealing with different doctors. Most of them tell you, they don’t talk to you. They don’t allow time for you to ask questions, or just like you had a video to show.” P-E3*

In addition to frequent and repeated use of the term “clear,” the feeling that the video was comprehensible was supported by comments such as “good pace,” “educational,” “enough info and easy to understand.” The concern about taVNS not being FDA approved was commonly expressed by participants across racial/ethnic groups.

The mean (SD) participant CEQ score was 7.2 (1.2) (median=7.2), with scores ranging from 5-9, indicating that all participants rated the credibility and expectancy as moderate to high. There were no differences between racial/ethnic groups in CEQ scores (p=0.57), and CEQ scores were not significantly correlated with MMS scores (r=0.06, p=0.83). Participant feedback was similar for both the full-length and segmented versions of the videos. One participant thought that the full-length version was too long to digest in a single viewing; additionally, the segmented versions seemed to encourage more participant interaction with the investigators between segments, and this enhanced engagement may facilitate rapport building and recruitment efforts.

### B. DEVELOPMENT of FINAL taVNS and TMS VIDEOS

#### B.1 taVNS Video Edits in Response to Feedback

The original plan was to re-shoot the videos in response to participant feedback. However, feedback was overwhelmingly positive, and thus the videos received only minor editing alterations. For example, due to the enhanced engagement of the shorter videos, we decided to only make segmented versions for the final taVNS and TMS video production. In addition, to allay participants’ concerns about FDA approval, we added the following to the narration of the taVNS Risks and Regulations video segment:

*“An invasive form of vagus nerve stimulation is FDA approved to treat epilepsy, depression, and stroke. Additional medical research is needed to gain FDA approval for the noninvasive form of TAVNS with these conditions and for other conditions like chronic pain.”*

#### B.2. TMS Video Development

Financial constraints were a major limiting factor, and we were only able to develop videos on TMS in English and Spanish due to the high costs associated with video production, conducting interviews and focus groups in Haitian-Creole, and for translation and transcription services. We applied lessons that were learned from the test video development for taVNS (i.e., preference for segmented versions and concerns about FDA approval), and the TMS content was divided into 4 short video segments entitled 1) Introduction (how TMS works), 2) TMS for Research, 3) TMS as a Treatment, and 4) Risks and Regulations.

The final videos are available for public use, and the web addresses for the taVNS and TMS videos can be found in **Table 3**.

**Table 3.**
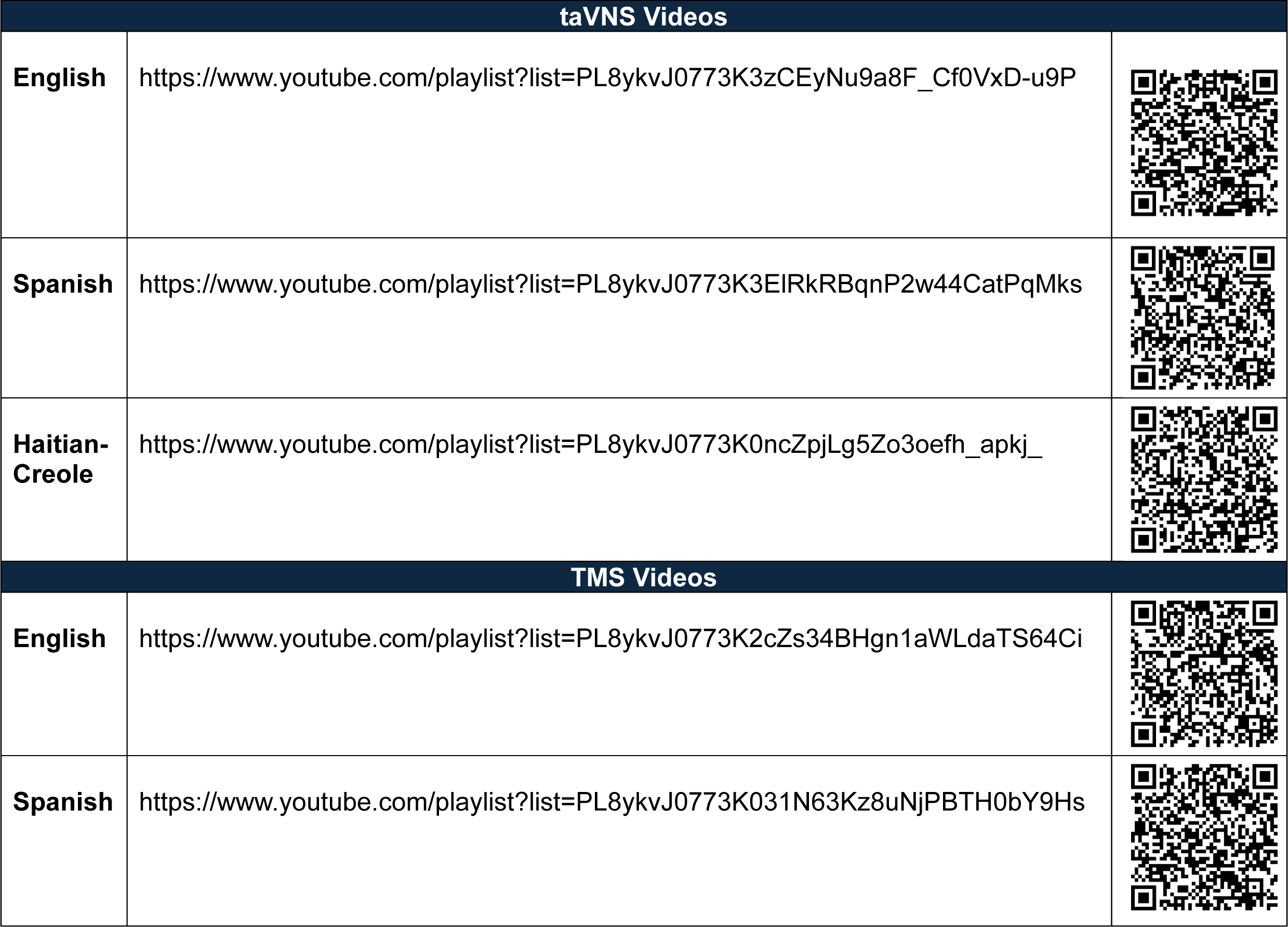
taVNS and TMS Video Links.

| taVNS Videos |  |
| --- | --- |
| **English** | https://www.youtube.com/playlist?list=PL8ykvJ0773K3zCEyNu9a8F_Cf0VxD-u9P |
| **Spanish** | https://www.youtube.com/playlist?list=PL8ykvJ0773K3EIRkRBqnP2w44CatPqMks |
| **Haitian-Creole** | https://www.youtube.com/playlist?list=PL8ykvJ0773K0ncZpjLg5Zo3oefh_apkj_ |
| TMS Videos |  |
| **English** | https://www.youtube.com/playlist?list=PL8ykvJ0773K2cZs34BHgn1aWLdaTS64Ci |
| **Spanish** | https://www.youtube.com/playlist?list=PL8ykvJ0773K031N63Kz8uNjPBTH0bY9Hs |

#### B.3. Final Video Feedback: Round 2

To get feedback on the final videos, we interviewed 3 new community members from each racial/ethnic group (**Table 2**). Since we did not have additional budget for translation services, we sought feedback from bilingual Spanish and Haitian-Creole speakers for the final round of interviews, and they watched the videos in their primary language but provided feedback in English. The bilingual Haitian-Creole speakers provided feedback on the final taVNS videos in Haitian-Creole and TMS video in English since we did not produce Haitian-Creole versions of the TMS videos. Participants in the second round of interviews had lower levels of mistrust that those in the first round, with mean MMS scores of 11.2 (3.2) (Median=12, range=6-15).

Overall, the videos for both taVNS and TMS videos were well received, and details of their feedback and preferences can be found in Supplement 1. For the taVNS videos, the mean (SD) participant CEQ score was 7.7 (1.1) (median=7.9), with scores ranging from 5.7-9, indicating that again all participants rated the credibility and expectancy as moderate to high. Once again, participants appreciated the clarity of the videos, calling them “clear”, “concise”, and “transparent.” In addition, word choice and translation were important to the Spanish speaking group. Specifically, they commented that although the translations used for the words “safety” and “seizure” were accurate, the literal translation was more alarming in Spanish.

After providing feedback on the videos for both taVNS and TMS, participants were asked if they were more interested in trying one of these interventions than the other. Key themes regarding treatment and learning preferences across groups were that they preferred the treatment option of TaVNS compared to TMS. All participants expressed greater interest in taVNS, and many indicated that there was a lack of clarity for how TMS would help with pain. For example, participants stated:

*“The first one you literally stated it could be effective with the chronic pain conditions. Especially, for me, the inflammation. Whereas the second one, it was more so for the smoking or the obsessive-compulsive disorder.” P-E7*

*“…Because the TMS helps with convulsions, depression, and things like that I didn’t feel it’s something that can help me with my chronic joint issue, with what I have.” P-S8*

This is also supported by the greater detail provided by participants in their descriptions of what they learned from the taVNS videos compared to the TMS videos. Many participants were able to discuss specific details on the theorized mechanisms for pain reduction via the vagus nerve with taVNS, but they could only vaguely discuss connections between brain stimulation and pain reduction with TMS.

Brochures of the same video content were also produced, and participants were asked to provide feedback on their preference for learning about taVNS and TMS from the videos or brochures. Surprisingly, there were varied preferences. All three Spanish speaking participants preferred learning from the video because you can see the procedure and participants undergoing the procedure, stating *“you see more of what will happen to you” P-S6,* and *“you see that the patient is calm…” P-S7*. However, two of the three Haitian-Creole speaking participants preferred the brochures over the videos. When providing reasons for their preference of the brochure over the video, the two participants discussed the importance of being able to take their time to read and research the information to discuss with their physician to make a decision about their care. For example:

*“I like the reading part. The video will be limited to me because even if it’s on TV or if you watch the video. But if I got it on my hand, I can read it, I can read it again, and then I can get that with me, and sit down somewhere, do more research…I keep the paper on my hand and I write it down. I notice word, which is something about what I don’t understand, I can go verify what’s the meaning of that…” P-C4*

*“You grab one, you read it, that’s how you are going to know the medication, that’s how when I go to the doctor’s I tell my doctor that I read the brochure outside and I see this medication is good for this certain thing.” P-C3*

### C. Study 2: Testing of the taVNS videos in a pilot randomized controlled trial

Twenty-eight people with peripheral neuropathy were recruited (**Figure 2**), of whom 14 received the taVNS videos in their preferred language (video group), and 14 received education via brochures only (control group). There were no differences between groups in participant demographics (i.e., race/ethnicity, sex) or medical condition (CIPN vs DN). The mean age of the participants was 58.2 (11.3), 71% were female, and 50% identified as Black and 50% identified as Hispanic/Latino. Additionally, sixty-one percent had CIPN, and 68% were using prescription medication to manage neuropathic symptoms, with gabapentin the most used medication (61%). On average, participants had high symptom burden with mean scores ranging from 6.1-8.0 on a scale of 0-10 for each of the following symptoms, pain, numbness, tingling, burning, and shooting/electric shocks.

**Figure 2.**
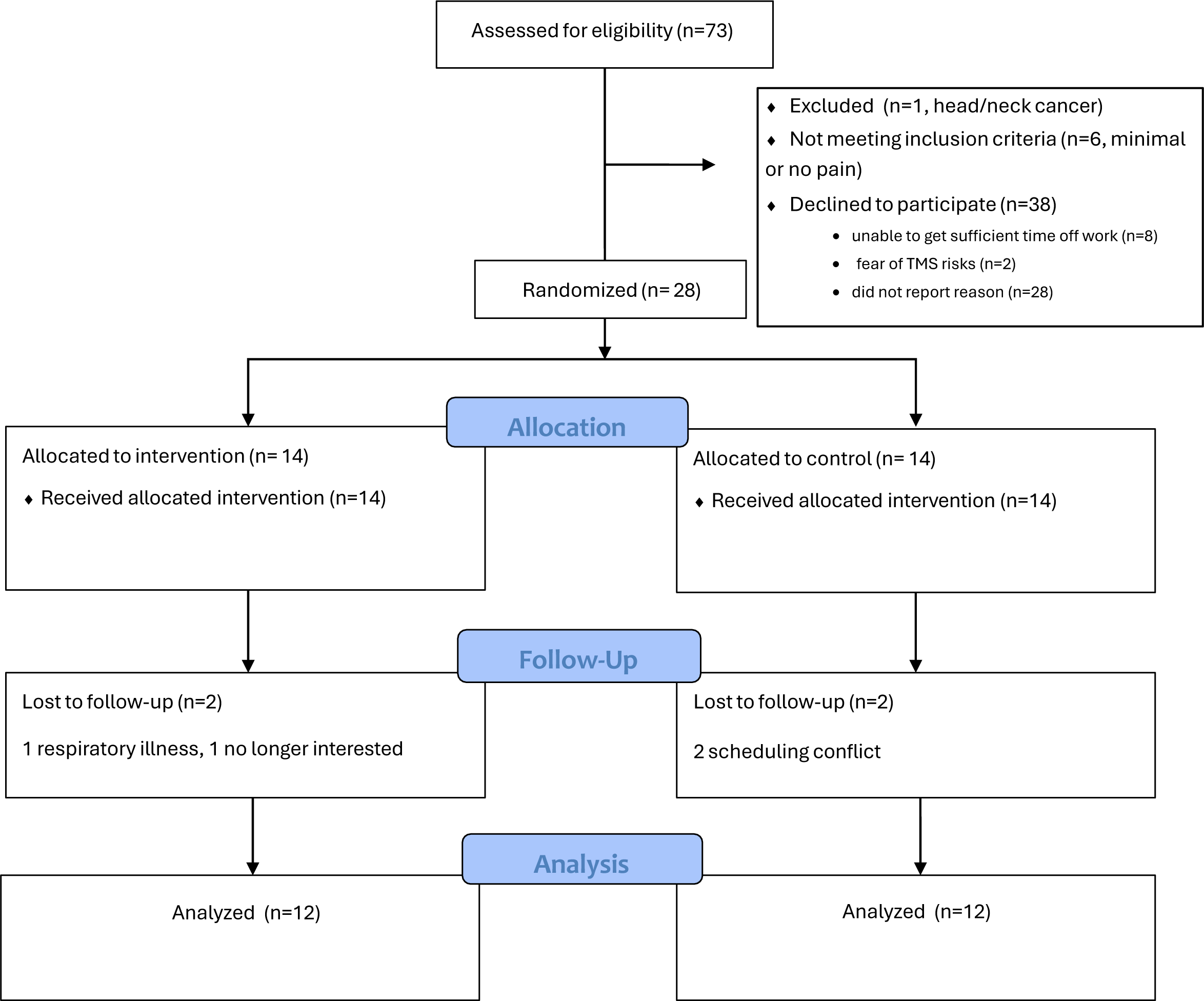
CONSORT Diagram.

After receiving education on taVNS, there was no meaningful difference in the median EXPECT scores between the video and control groups, with respective mean values of 8.0 and 7.5 (median values of 8.2 for both groups, and 95% confidence intervals for the means of 7.2-8.7 and 6.4-8.7, p=0.8). However, the video exposed group had less variability in EXPECT scores, with a standard deviation of 1.3 compared to 2.0 for the control group (**Figure 3**). Similarly, there were no differences between groups after completing the trial, with mean EXPECT scores at visit 3 of 8.1 (0.9) and 8.2 (1.9) for the video and control groups (median values of 8.1 and 9.0 and 95% confidence intervals for the means of 7.6-8.7 and 6.9-9.4, p=0.6), respectively. Further, there was no significant change between visit 1 and visit 3 EXPECT scores (mean difference=-0.2, p=0.28).

**Figure 3.**
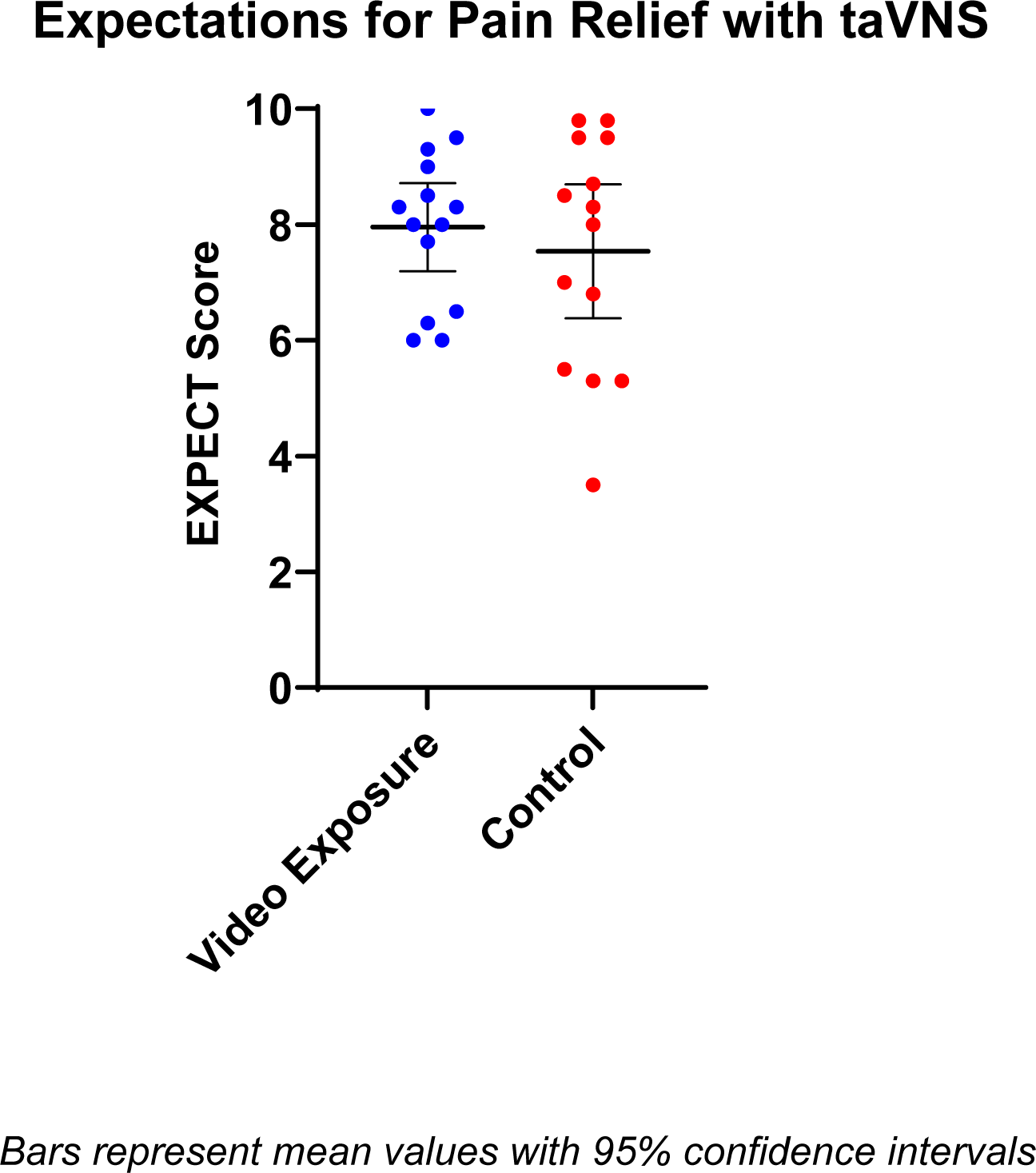
Group Differences in EXPECT Scores.

## DISCUSSION

In these 2 studies, we used community participatory research principles to develop and test culturally sensitive informational videos on taVNS and TMS. These videos were designed specifically to enhance the recruiting and consenting processes for taVNS and TMS research with Black English-speaking, Hispanic/Latino Spanish-speaking, and Haitian-Creole-speaking people by providing the information in an easily digestible format, with input from the target communities, and with representation of these communities within the videos. The final video products were well received and generated interest in these modalities among the participants. We anticipated that the videos would increase expectancy for pain relief with taVNS, but there were no differences in EXPECT scores between those who viewed the videos and those who learned about taVNS from brochures only. Although unanticipated, this finding likely increases the potential value of these videos for use in research. An undue increase in expectations could be a potential confounding factor in studies, and our results suggest that these videos are neutral and do not generate unrealistic or misleading expectations for pain relief with taVNS.

A strength of this study was the range of medical mistrust observed in the sample, with scores from the lowest possible to the highest possible on the MMS. In a racially and ethnically diverse sample of 615 American adults and adolescents the mean MMS score was 13.3,^20^ which is lower than the mean score of 15.8 found in our sample. Much has been made of medical mistrust as a key barrier to recruitment and retention of minoritized groups in research; however, in this study high medical mistrust did not result in low perceived credibility, expectancy, or interest in participating in taVNS research. Feelings of uncertainty are known to contribute to minoritized patients’ reluctance to participate in research, and poor quality information can contribute to participants’ uncertainty.^29^ Research on Media Richness Theory has shown that richer media are viewed as more credible, and that a video with audio medium will be perceived as higher in credibility than a picture with text medium.^16, 17^ Therefore, it is plausible that the high CEQ scores and expressed interest in taVNS research observed in our study was a result of the videos mitigating feelings of uncertainty. However, additional research is needed to confirm this.

It was interesting that all community participants in round 2 of Study 1 expressed greater interest in taVNS than TMS, and they demonstrated greater understanding of the proposed mechanisms for pain relief with taVNS compared to TMS. Therefore, in this sample of chronic pain patients, understanding treatment mechanisms appeared to be an important factor in treatment preference across cultural/language groups. There were also group differences in their preference for learning material format, with only members of the Haitian-Creole speaking group expressing a preference for brochures over video. It is well known that Haitian immigrants have complex language and cultural barriers that limit access to healthcare services.^30-32^ Haitian participants in this study differed from the Black English-speaking and Hispanic/Latino Spanish-speaking participants in that they preferred brochures over videos for learning about NIBS, and this highlights the importance of using nuanced approaches to optimize community engagement in research. We plan to build on these lessons in the future by providing future potential Creole-speaking participants with brochures in both Haitian Creole and in English, so that they will have tools to discuss the studies with their families and providers.

Although these studies achieved their aims, there are key limitations worth noting. The qualitative analyses were done on translated transcripts of the Spanish and Haitian-Creole audio recordings rather than transcripts in the native languages. Additionally, no Haitian-Creole speakers were involved with analyzing the transcripts. It is possible that salient points were missed or misinterpreted during translation, but this risk was mitigated by member checking during CAB meetings. It is also important to note that while many of the participants were originally from different countries or different parts of the United States, all but one were current residents of the Miami-Dade County area. Thus, our findings may not be reflective of all other communities, and researchers should consider the nuances of their respective communities and possibly test the videos before implementing them.

Due to budgetary constraints and the high costs associated with translation services, we were unable to produce Haitian-Creole versions of the TMS videos. Outside of costs associated with video production and personnel for conducting the interviews, over $17,000 was spent on translation and transcription services across both studies, with more funds dedicated to Haitian-Creole translation than either English or Spanish, despite not including Haitian-Creole speakers in study 2. For example, the rates that we received for transcribing the audio recordings of the interviews varied greatly depending on the language, with the per recorded minute rate at $1.50 for English-to-English, $5.50 for Spanish-to-English, and $20 per audio minute for Haitian-Creole-to-English. This 13-fold increase in transcription costs for Haitian-Creole audio is a significant barrier to including this underserved population in health research opportunities. Further, compared to the English and Spanish recordings, the transcription service used was more likely to deem the audio quality of the Haitian-Creole interviews as “difficult” which added $0.50 per minute to the cost.

Our goal was to utilize participatory research principles in this research, and we implemented this by using the CAB and seeking input from community members. Ideally, we would have used a more mutually collaborative model, in which community members have input throughout the research process, including conceptualization, data collection, and dissemination of the findings. However, these studies were the first steps in developing infrastructure for a research agenda focused on the use of NIBS for equitable pain management; thus, we did not have strong existing relationships with community partners, and most of our participants were not experienced with research involvement. Additionally, community participants expressed difficulty generating unguided input due to the novelty of taVNS. As a result, we chose to use a technical-scientific and positivist^18^ in which community members were only used as a sounding board for feedback.

Together, these 2 studies describe the process of engaging stakeholders to develop and test culturally sensitive informational videos on taVNS and TMS. The iterative process used to develop the videos in this project resulted in enhanced community awareness, engagement, and interest in our research agenda. Our hope is that the videos produced in this project will provide NIBS researchers with culturally sensitive and useful tools to engage minoritized communities in their research.

## Data Availability

All data produced in the present study are available upon reasonable request to the authors

## Acknowledgements

We would like to thank April Mann for her contributions to editing, the University of Miami Behavioral and Community-Based Research Shared Resource for their assistance with participant recruitment, and Peter Fried for his consultant role.

## Authorship Contributions

As the corresponding author, Marlon L. Wong was responsible for conceptualization, data curation, formal analysis, funding acquisition, investigation, methodology, project administration, resources, supervision, validation, visualization, and writing.

Lisa M. McTeague made substantial contributions to the conceptualization, funding acquisition, investigation, resources, supervision, and writing.

Chelsea Miller made substantial contributions to the conceptualization, data curation, formal analysis, methodology, validation, visualization, and writing.

Gabriel Gonzalez made substantial contributions to the data curation, formal analysis, investigation, methodology, and writing.

Melissa M. Tovin made substantial contributions to the conceptualization, funding acquisition, methodology, visualization, and writing.

Frank J. Penedo made substantial contributions to the conceptualization, funding acquisition, methodology, and writing.

Eva Widerstrom-Noga made substantial contributions to the conceptualization, funding acquisition, supervision, and writing.

---

**Supplement 1.**
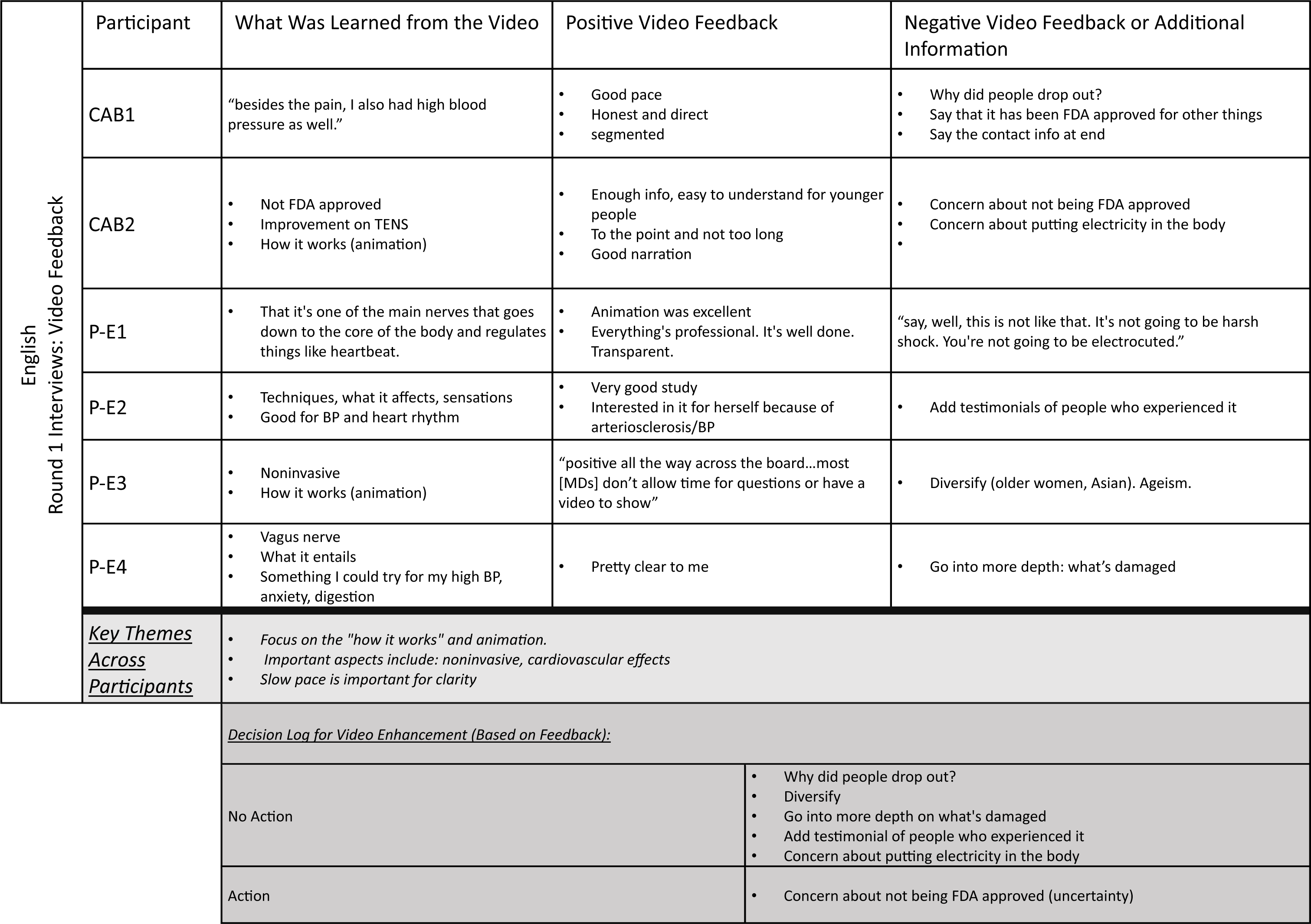
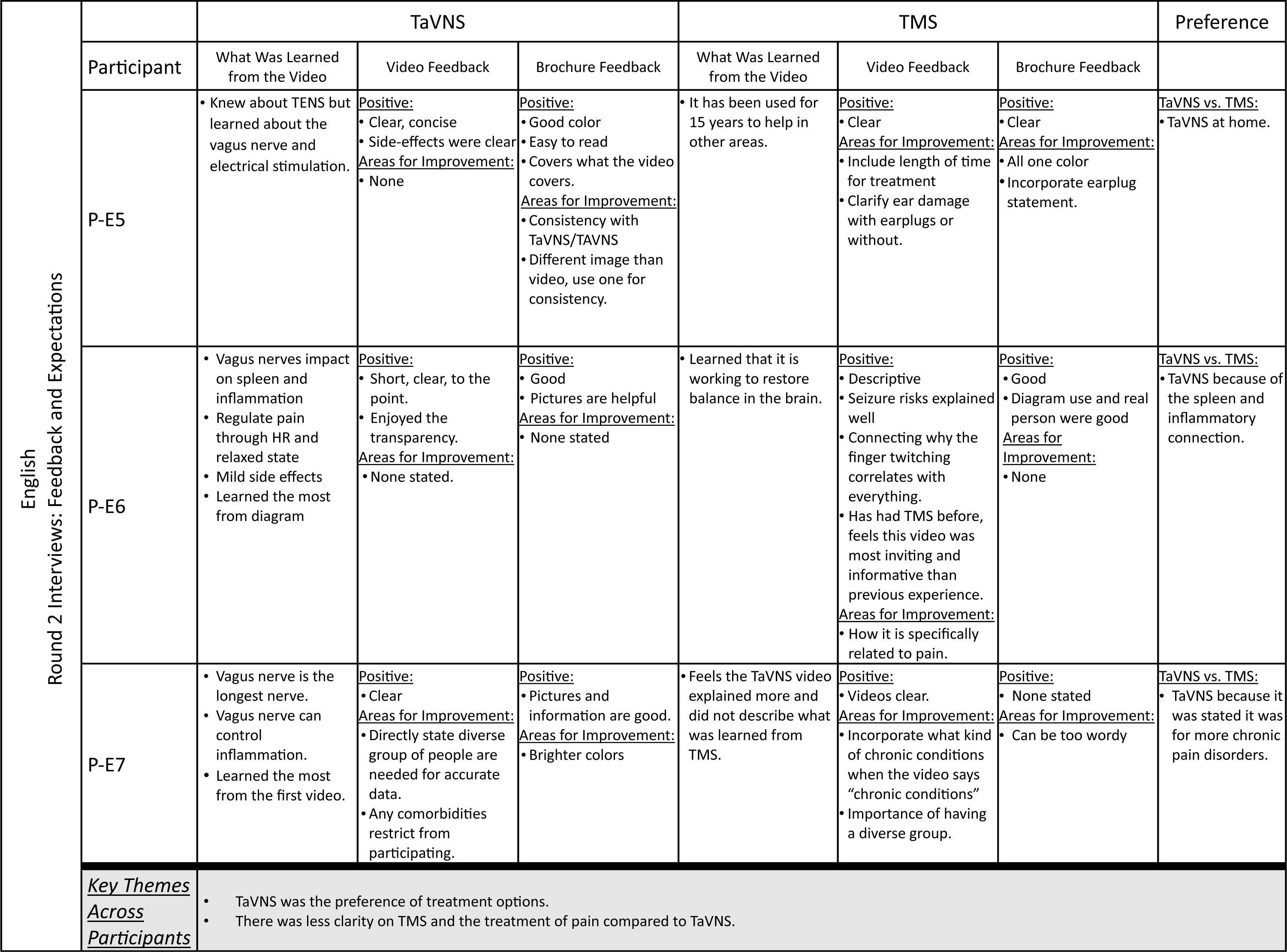
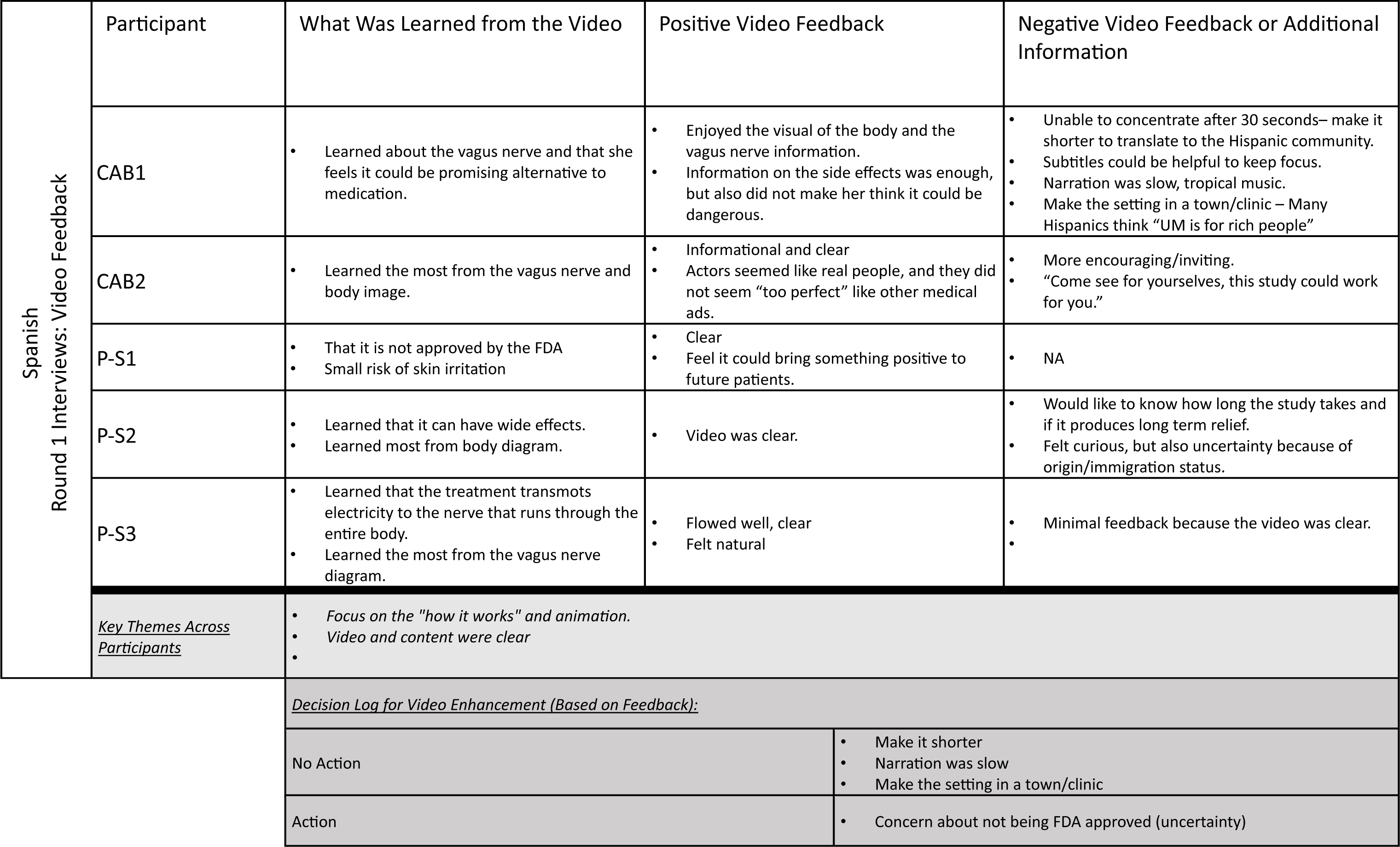
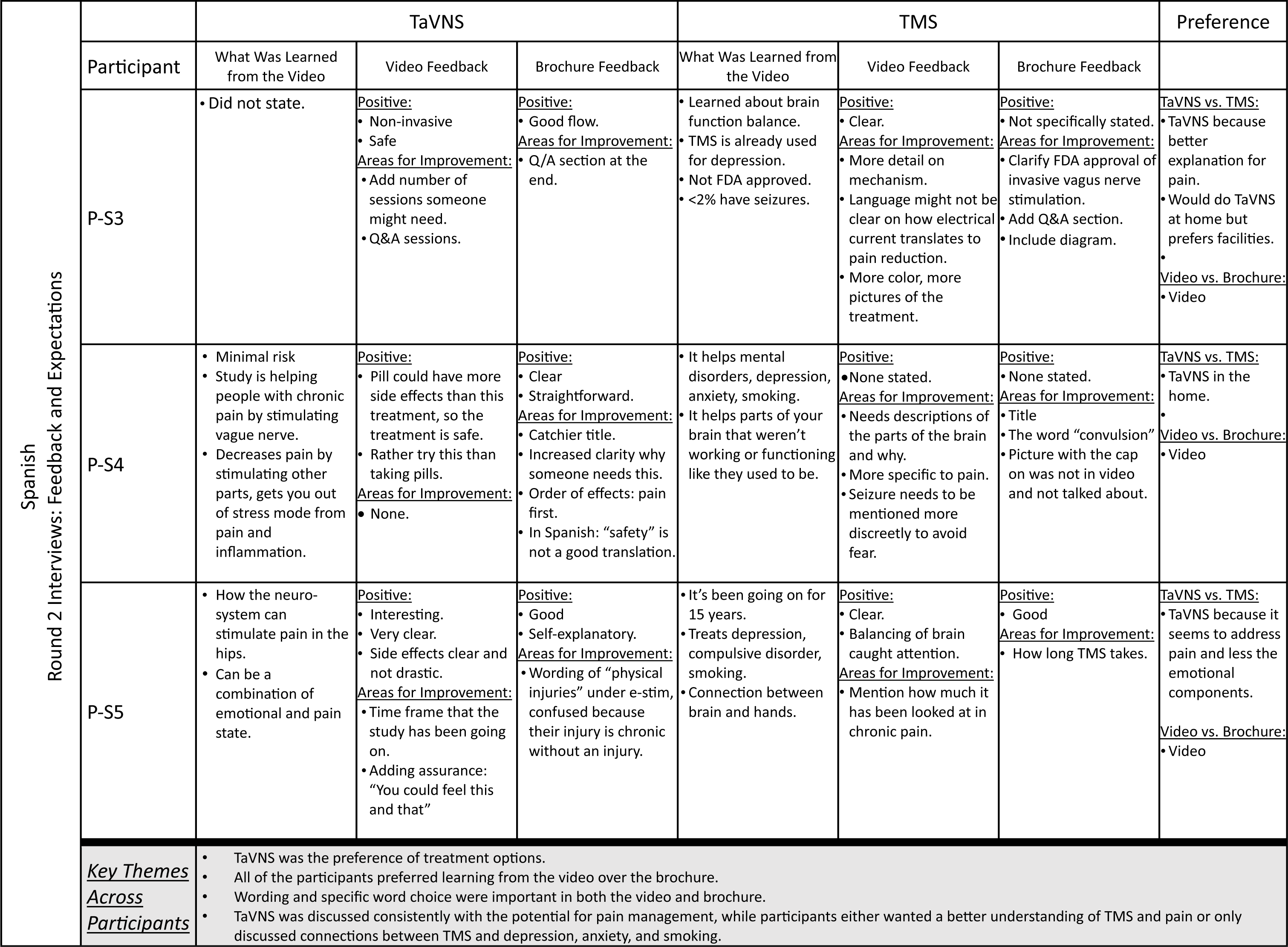
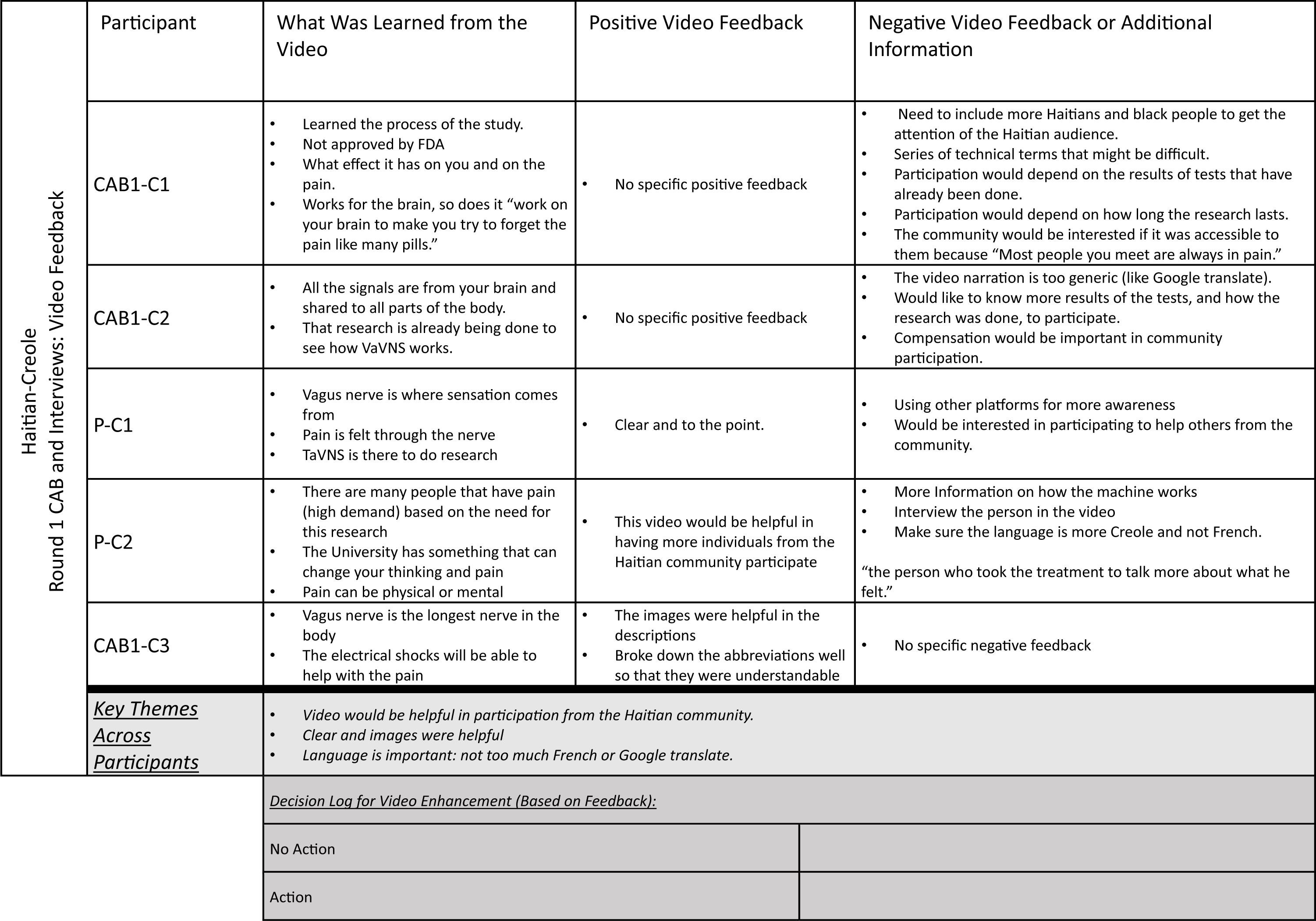
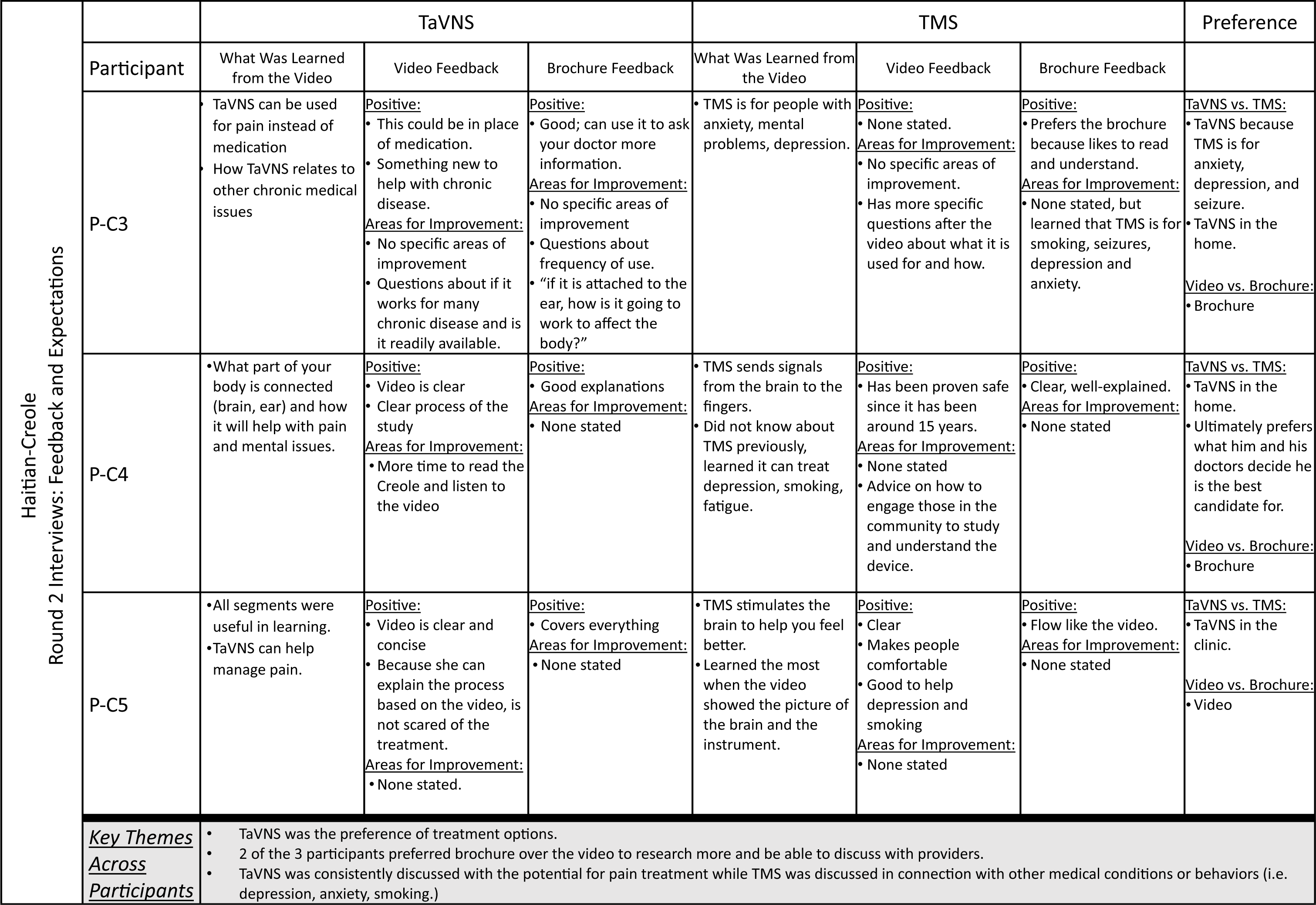
Qualitative Data Matrices.

